# Investigation of ceftriaxone-resistant *Neisseria gonorrhoeae* detected in Scotland, 2018-2024

**DOI:** 10.1101/2024.10.24.24315890

**Authors:** Martin P McHugh, Kirsty Aburajab, Alexandra Maxwell, John Anderson, Fraser Cairns, Seb Cotton, Andrew Gough, Brian Malloy, Katharine Mathers, Lynne Renwick, Jill Shepherd, Kate E Templeton

**Affiliations:** NHS Lothian, Edinburgh, United Kingdom; Edinburgh Medical School, University of Edinburgh, Edinburgh, United Kingdom; School of Medicine, University of St Andrews, St Andrews, United Kingdom; NHS Forth Valley, Falkirk, United Kingdom; NHS Greater Glasgow and Clyde, Glasgow, United Kingdom

## Abstract

**Objectives:** Describe the clinical, phenotypic, and genomic characteristics of ceftriaxone-resistance *Neisseria gonorrhoeae* from Scotland

**Methods:** Cases were identified in routine care from 2018-2024. Minimum inhibitory concentrations were determined for seven antimicrobial agents. Whole genome sequencing was performed with Illumina and Oxford Nanopore Technology instruments. A phylogeny containing global ceftriaxone-resistant genomes was generated with Parsnp. A collection of ST8780 genomes was also analysed to give further context using reference based mapping with Snippy.

**Results:** There were five cases of ceftriaxone-resistant *N. gonorrhoeae* detected. One case (MLST ST1903) clustered within the FC428 lineage in a returning traveller from an Asia-Pacific country. Two cases belonged to the recently described extensively drug resistant MLST ST16406, a returning traveller from an Asia-Pacific country and a sexual contact within Scotland. The final two cases were a resident of an Asia-Pacific country and a sexual contact within Scotland, both belonged to MLST ST8780. These were distinct from other publicly available ST8780 genomes suggesting a novel introduction of the mosaic *penA* 60.001 allele. All cases were initially treated with ceftriaxone-based regimes, four returned for test of cure and showed clearance of infection.

**Conclusions:** As ceftriaxone resistance is increasingly identified, multiple public health interventions are required to reduce the impact of resistance on gonorrhoea treatment globally.

## INTRODUCTION

*Neisseria gonorrhoeae* is a leading cause of sexually transmitted infections globally. WHO includes *N. gonorrhoeae* on the list of Priority Pathogens due to significant antimicrobial resistance (AMR), reflecting the significant global health challenge posed by this pathogen [1]. Currently, ceftriaxone is the backbone of gonorrhoeae treatment. Ceftriaxone resistance has been increasingly identified particularly associated with countries in the Asia-Pacific region, and in recent years extensively drug-resistant (XDR) cases with few treatment options have been described [2–5].

We describe the identification, clinical management, laboratory, and genomic investigation of five ceftriaxone-resistant *N. gonorrhoeae* isolated in Scotland.

## MATERIALS AND METHODS

This study describes five cases of ceftriaxone-resistant *N. gonorrhoeae* identified in Scotland as part of routine surveillance undertaken by the Scottish Bacterial Sexually Transmitted Infections Reference Laboratory between 2018-2024. All *N. gonorrhoeae* isolates were submitted by regional diagnostic laboratories for antimicrobial sensitivity testing (AST) and further characterisation. Species identification was confirmed with biochemical testing and MALDI-TOF (Bruker). AST was performed by agar incorporation and Etest strips (BioMerieux), minimum inhibitory concentrations (MICs) were interpreted with EUCAST v14.0 breakpoints. Genomic DNA was extracted from culture suspensions, short read libraries were prepared with the Illumina DNA Prep kit and sequenced on a MiSeq or NextSeq 2000 (Illumina). Short reads were trimmed with Trimmomatic v0.39 and assembled with SPAdes v3.15.5 (with the --isolate and -- cov-cutoff 15 options). PyngoST was used to perform multilocus sequencing typing (MLST) and *N. gonorrhoeae* sequence typing for antimicrobial resistance (NG-STAR). Resistance markers were identified from assemblies using PathogenWatch. Published sequences from ceftriaxone-resistant *N. gonorrhoeae* were used for global context (Table S1), a core genome alignment was generated and recombination masked with Parsnp v1.5.3, and the phylogeny visualised with iTOL v6. To further investigate ST8780, short reads were downloaded from the NCBI Short Read Archive (Table S2), trimmed, mapped to the WHO O reference genome (Accession NZ_CP145037.1) with Snippy v4.6.0, recombination masked with Gubbins v2.4.1, and the resulting phylogeny visualised with iTOL. Long reads were generated for SGC-24-001 and SGC-24-002 using the Rapid Barcoding v14 library preparation kit and an R10.4.1 flowcell on a GridION sequencer with super high accuracy basecalling in MinKNOW v24.02.16. Long reads were quality filtered with Filtlong v0.2.1 and assemblies generated with Flye v2.9.3 (with the -- nano-hq and --genome-size 2.3m options). Resulting assemblies were polished with Medaka v1.11.3, Polypolish v0.6.0, and Pypolca 0.3.1. Patient demographics and treatment histories were retrieved from electronic health records.

## RESULTS

### Case 1, SGC-23-001

Case 1 (SGC-23-001) was a male who presented to sexual health services in 2018 with urethral discharge after unprotected sex in an Asia-Pacific country (Table S3). Urine was NAAT positive for *N. gonorrhoeae* and a urethral swab grew *N. gonorrhoeae*. The patient was given a 500mg intramuscular dose of ceftriaxone and 1g oral azithromycin as per national treatment guidance at the time. Fourteen days after empirical treatment AST results were reported which showed multidrug resistance including ceftriaxone (MIC 0.75 mg/L) and cefixime (MIC 2.0 mg/L), but not azithromycin (MIC 0.25 mg/L) and the patient was given a further 2g oral dose of azithromycin (Table S4). Test of cure (TOC) NAAT was negative. No sexual contacts within Scotland were identified.

### Case 2, SGC-23-002

The second case (SGC-23-002) was a male who presented to sexual health services in 2022 with a two week history of urethral discharge and dysuria after unprotected sex in an Asia-Pacific country (Table S3). The urine NAAT was positive, and *N. gonorrhoeae* was grown from a urethral swab. Intramuscular ceftriaxone (1g) was given, and the patient had a negative TOC two weeks later. AST identified this as an XDR strain with non-susceptibility to six of the seven tested agents, including ceftriaxone (MIC 0.19 mg/L), cefixime (2.0 mg/L), and high level resistance to azithromycin (MIC >256.0 mg/L; Table S4).

### Case 3, SGC-23-003

Case 3 (SGC-23-003) was a female sexual contact of case 2 in Scotland, case 2 and 3 reported no other recent sexual contacts (Table S3). The patient reported two days of abdominal pain, on examination profuse discharge was found but no evidence of pelvic inflammatory disease. A vaginal swab was NAAT positive and *N. gonorrhoeae* was isolated from a cervical swab. Intramuscular ceftriaxone (1g) was given, and the patient had a negative TOC two weeks later. AST results were similar to case 2 although the ceftriaxone MIC was slightly higher (MIC 0.38 mg/L; Table S4).

### Case 4, SGC-24-002

Case 4 (SGC-24-002) was an asymptomatic female who tested NAAT positive on a vaginal swab after unprotected sex in 2024 (Table S3). The patient was treated with intramuscular ceftriaxone (1g) and was negative at TOC. AST showed multidrug resistance including ceftriaxone (MIC 0.75 mg/L) and cefixime (MIC 2.0 mg/L; Table S4). Only one sexual partner was identified.

### Case 5, SGC-24-001

Case 5 (SGC-24-001) was the male sexual contact of case 4 in Scotland. Case 5 reported two other female sexual contacts, but these individuals could not be traced for testing (Table S3). Case 5 reported dysuria and urethral discharge for one week prior to presentation. The urine NAAT was positive, and *N. gonorrhoeae* was grown from a urethral swab. AST showed the same multidrug resistance as case 4 with slightly higher cephalosporin MICs (ceftriaxone MIC 1.0 mg/L and cefixime MIC 3.0 mg/L; Table S4). Intramuscular ceftriaxone (1g) was given at presentation, when culture results were reported the patient was further treated with intramuscular gentamicin (240mg) and oral azithromycin (2g). The patient was a resident of an Asia-Pacific country and moved back before TOC was performed, further follow-up has been unsuccessful.

### Genomic investigation

Genotypic mechanisms conferring all detected phenotypic resistance were identified in the genomes, including the mosaic *penA*-60.001 conferring cephalosporin resistance, *gyrA* and *parC* mutations conferring ciprofloxacin resistance, and the *rpsJ* V57M polymorphism and *tetM* conferring tetracycline resistance (Table S5). The isolates belonged to diverse sequence types which suggested distinct origins. Phylogenetic analysis with a background of international ceftriaxone-resistant isolates revealed that SGC-23-001 clustered within a diverse clade containing MLST ST1903 and related STs identified in 2015-2021 from China and five other countries (Figure 1). SGC-23-001 was also assigned to NG-STAR ST233 which was common in this clade, indicating the presence of similar AMR mechanisms. The closest match in this dataset was the FC428 type strain from Japan in 2015 (428 single nucleotide polymorphisms [SNPs]). SGC-23-002 and SGC-23-003 differed from each other by two SNPs, consistent with direct transmission as supported by partner notification. Additionally, the two genomes differed by a median of 4 SNPs (range 1-7 SNPs) to three other MLST ST16406 and NG-STAR ST4465 genomes identified in Austria, England, and France in 2022 (Figure 1). High-level azithromycin resistance was conferred by the 23S rDNA A2059G mutation (*Escherichia coli* numbering). No SNPs were identified between SGC-24-001 and SGC-24-002 which supports direct transmission. Additionally, these two genomes were distinct in this analysis differing to the nearest genome (GD2021236 from China in 2021) by 3430 SNPs. SGC-24-001 and SGC-24-002 were assigned to MLST ST8780 and NG-STAR ST5998, this was a novel NG-STAR ST indicating these genomes are unique within the global database. To further investigate the origins of SGC-24-001 and SGC-24-002, a collection of MLST ST8780 genomes were identified and a phylogeny generated (Figure S1). The Scottish isolates were the only ones containing the mosaic *penA*-60.001 allele, indicating that this is a novel acquisition of ceftriaxone resistance in this ST. The Scottish isolates were most closely related to an isolate from Thailand in 2018 (949 SNPs). SGC-24-001 and SGC-24-002 carried the cryptic plasmid, conjugative plasmid, and the African-type β-lactamase (*bla*_TEM-1_) plasmid.

**Figure 1.**
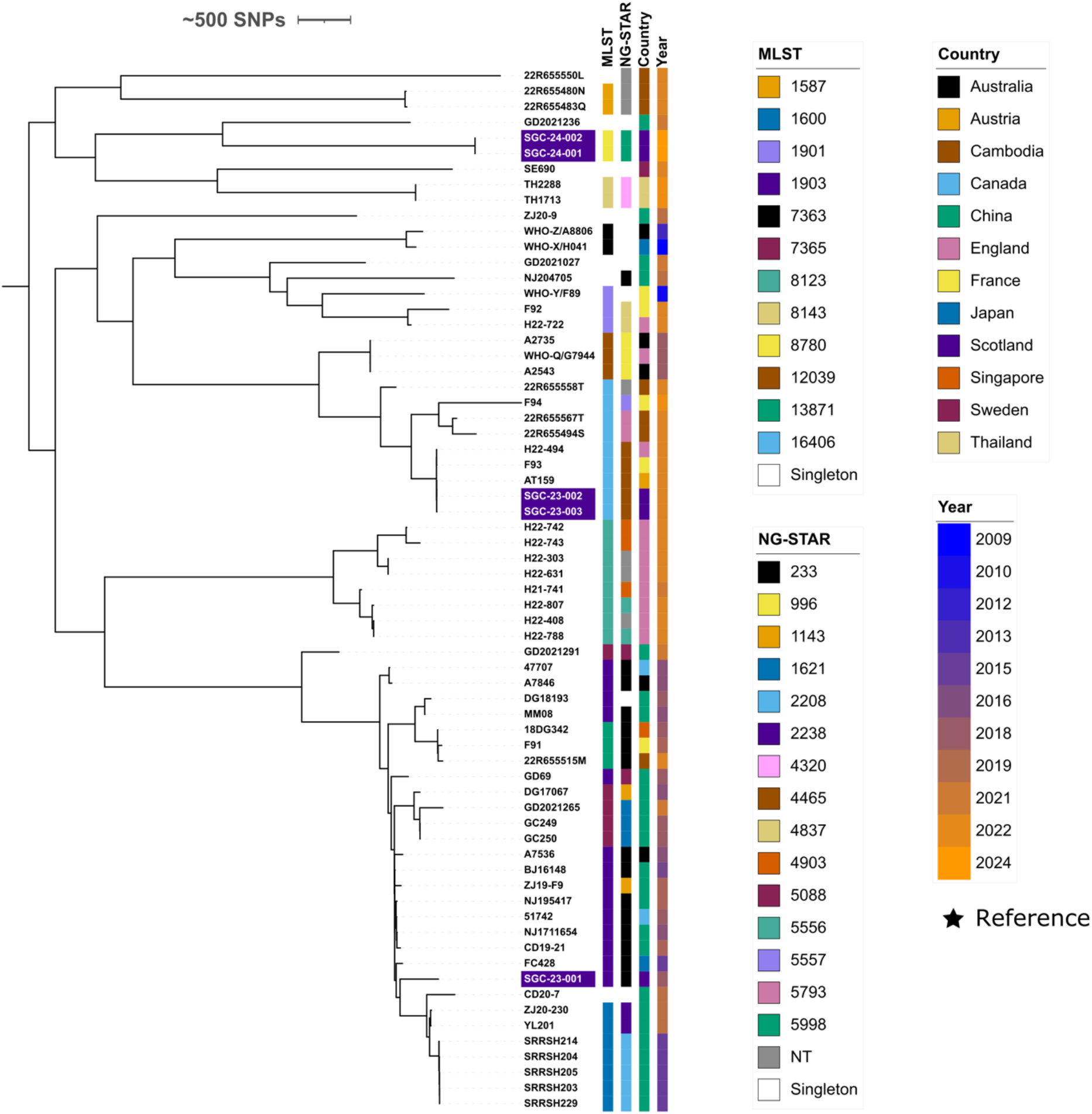
Maximum-likelihood phylogeny of five Scottish and 62 international ceftriaxone-resistant *N. gonorrhoeae*. Scottish genomes are highlighted in purple. Metadata bars coloured as per figure legend.

## DISCUSSION

We describe five cases of ceftriaxone-resistant *N. gonorrhoeae* acquired in, or involving residents of, Asia-Pacific countries identified in Scotland. Case 1 from 2018 belonged to the FC428-related clone (MLST ST1903) which has acquired the mosaic *penA* allele and is frequently identified in the Asia-Pacific [4]. Cases 2 and 3 from 2022 belonged to ST16406 and were resistant to current first-line antimicrobial agents ceftriaxone, azithromycin, and ciprofloxacin. ST16406 was first described in 2022 and has been identified in travellers returning from the Asia-Pacific region to Austria, England, and France [2,3,5]. We identified a median of 4 SNPs between cases identified in Austria, England, France, and Scotland which reflects the short collection time (all in 2022) but also suggests a rapidly spreading clone. To our knowledge, cases 4 and 5 are the first ST8780 identified with ceftriaxone resistance. ST8780 belongs to the generally antimicrobial-susceptible genomic lineage B of *N. gonorrhoeae*, indicating that mosaic *penA* 60.001 is spreading further within the global population as recently identified by Golparian *et al* [6]. Concerningly, Case 5 reported two sexual contacts who could not be traced and may lead to further ceftriaxone-resistant cases within Scotland.

Four patients were successfully treated with ceftriaxone-based regimes despite *in vitro* resistance, the remaining patient did not return for TOC. There is growing evidence that the ceftriaxone resistance breakpoint is too low for genital infections, although treatment success at other sites such as the oropharynx need to be considered in any breakpoint change [7–9]. We also identified different cephalosporin MICs within two transmission pairs, although the reported values are within the expected variability of the method (± two doubling dilutions). Our reliance on cultured isolates is a potential limitation as ∼50% of gonorrhoea cases in Scotland are diagnosed by NAAT only and no isolate is available. It is possible that culture-negative ceftriaxone resistant cases have occurred that would not be included in our study. To reduce the impact of this, patients that remain symptomatic after treatment are reviewed to ascertain if ceftriaxone resistance may be present.

## CONCLUSION

Resistance to ceftriaxone poses a significant risk to *N. gonorrhoeae* treatment guidelines globally, although in the cases described here who had TOC first line agents were successful. Efforts to expand access to rapid molecular diagnostics to determine sensitivity to ceftriaxone and alternative agents are required to reduce the selection of resistance, and the introduction of novel therapeutic agents must be carefully managed and combined with robust surveillance to identify emerging resistance. The 4CMenB vaccine has been approved in the UK for individuals at high risk of gonorrhoea, the effect of this vaccine on infections and AMR should be closely monitored. Finally, sustained messaging to reduce STI transmission both domestically and in international travellers should be prioritised.

## ETHICS STATEMENT

Ethical approval for this work was granted by the Lothian National Research Scotland BioResource (Reference 20/ES/0061)

## DATA AVAILABILITY STATEMENT

Sequence data has been uploaded to the European Nucleotide Archive (BioProject PRJEB73456)

## ACKNOWLEDGEMENTS

We would like to thank Matthew Holden, Sharif Shaaban, and Lesley Wallace for providing feedback on the manuscript. We thank diagnostic microbiology laboratories for providing isolates for characterisation.

## CONTRIBUTIONS

MPM: conceptualisation, methodology, data curation, investigation, writing – original draft, guarantor; KA, AM, JA, FC, SC, AG, BM, KM, LR and JS: data curation, writing – review and editing; KET: conceptualisation, writing – review and editing

