## Supplemental material for "Investigation of ceftriaxone-resistant *Neisseria gonorrhoeae* detected in Scotland, 2018-2024"

**Table S1.** Ceftriaxone resistant genomes used in Figure 1

| Sample | BioProject | BioSample | Country | Year | MLST | NG-STAR |
|---|---|---|---|---|---|---|
| 47707 | PRJNA415047 | SAMN07813261 | Canada | 2017 | 1903 | 233 |
| 51742 | PRJNA526031 | SAMN11082624 | Canada | 2018 | 1903 | 233 |
| 18DG342 | PRJNA508549 | SAMN10524911 | Singapore | 2018 | 13871 | 233 |
| A2543 | PRJEB29480 | SAMEA5061681 | Australia | 2018 | 12039 | 996 |
| A2735 | PRJEB29480 | SAMEA5061682 | Australia | 2018 | 12039 | 996 |
| A7536 | PRJNA416507 | SAMN07958489 | Australia | 2017 | 1903 | 233 |
| A7846 | PRJNA416507 | SAMN07958490 | Australia | 2017 | 1903 | 233 |
| AT159 | PRJNA839941 | SAMN28560617 | Austria | 2022 | 16406 | 4465 |
| BJ16148 | PRJNA614605 | SAMN14433815 | China | 2016 | 1903 | 233 |
| CD19-21 | PRJNA560592 | SAMN33016850 | China | 2019 | 1903 | 233 |
| CD20-7 | PRJNA560592 | SAMN33016861 | China | 2020 | 7356 | 4906 |
| DG17067 | PRJNA874857 | SAMN30571139 | China | 2017 | 7365 | 1143 |
| DG18193 | PRJNA778600 | SAMN22965531 | China | 2018 | 1903 | 3859 |
| F91 | PRJNA554496 | SAMN12321478 | France | 2019 | 13871 | 233 |
| F92 | PRJNA901436 | SAMN31713390 | France | 2022 | 1901 | 4837 |
| F93 | PRJNA998219 | SAMN36700584 | France | 2022 | 16406 | 4465 |
| F94 | PRJNA998219 | SAMN36700585 | France | 2023 | 16406 | 5557 |
| FC428 | PRJNA416507 | SAMN07958491 | Japan | 2015 | 1903 | 233 |
| GC249 | PRJNA560592 | SAMN13221485 | China | 2018 | 7365 | 1621 |
| GC250 | PRJNA560592 | SAMN13221486 | China | 2018 | 7365 | 1621 |
| GD2021027 | PRJNA957547 | SAMN34257423 | China | 2021 | 10314 | 5555 |
| GD2021236 | PRJNA957547 | SAMN34257424 | China | 2021 | 1588 | 5554 |
| GD2021265 | PRJNA957547 | SAMN34257425 | China | 2021 | 7365 | 1621 |
| GD2021291 | PRJNA957547 | SAMN34257434 | China | 2021 | 7365 | 5088 |
| GD69 | PRJNA874857 | SAMN30571138 | China | 2018 | 1903 | 5088 |
| H21-741 | PRJEB57389 | SAMEA112168211 | England, UK | 2021 | 8123 | 4903 |
| H22-303 | PRJEB57389 | SAMEA112168215 | England, UK | 2022 | 8123 | - |
| H22-408 | PRJEB57389 | SAMEA112168217 | England, UK | 2022 | 8123 | - |
| H22-494 | PRJEB57389 | SAMEA112168220 | England, UK | 2022 | 16406 | 4465 |
| H22-631 | PRJEB57389 | SAMEA112168214 | England, UK | 2022 | 8123 | - |
| H22-722 | PRJEB57389 | SAMEA112168219 | England, UK | 2022 | 1901 | 4837 |

| | | | | | | |
|---|---|---|---|---|---|---|
| H22-742 | PRJEB57389 | SAMEA112168212 | England, UK | 2022 | 8123 | 4903 |
| H22-743 | PRJEB57389 | SAMEA112168213 | England, UK | 2022 | 8123 | 4903 |
| H22-788 | PRJEB57389 | SAMEA112168218 | England, UK | 2022 | 8123 | 5556 |
| H22-807 | PRJEB57389 | SAMEA112168216 | England, UK | 2022 | 8123 | 5556 |
| MM08 | PRJNA778600 | SAMN22965526 | China | 2017 | 1903 | 233 |
| NJ1711654 | PRJNA224116 | SAMN12252293 | China | 2017 | 1903 | 233 |
| NJ195417 | PRJNA916595 | SAMN32487317 | China | 2019 | 1903 | 233 |
| NJ204705 | PRJNA916595 | SAMN32487324 | China | 2020 | 11710 | 233 |
| SE690 | PRJNA924675 | SAMN32765978 | Sweden | 2022 | 8130 | 4859 |
| SGC-23-001* | PRJEB73456 | SAMEA115360394 | Scotland, UK | 2018 | 1903 | 233 |
| SGC-23-002* | PRJEB73456 | SAMEA115360395 | Scotland, UK | 2022 | 16406 | 4465 |
| SGC-23-003* | PRJEB73456 | SAMEA115360396 | Scotland, UK | 2022 | 16406 | 4465 |
| SGC-24-001* | PRJEB73456 | SAMEA116302675 | Scotland, UK | 2024 | 8780 | 5998 |
| SGC-24-002* | PRJEB73456 | SAMEA116302676 | Scotland, UK | 2024 | 8780 | 5998 |
| SRRSH203 | PRJNA606927 | SAMN14116987 | China | 2015-17 | 1600 | 2208 |
| SRRSH204 | PRJNA606927 | SAMN14116988 | China | 2015-17 | 1600 | 2208 |
| SRRSH205 | PRJNA606927 | SAMN14116989 | China | 2015-17 | 1600 | 2208 |
| SRRSH214 | PRJNA606927 | SAMN14116991 | China | 2015-17 | 1600 | 2208 |
| SRRSH229 | PRJNA606927 | SAMN14116992 | China | 2015-17 | 1600 | 2208 |
| WHO-Q/G7944 | PRJEB26560 | SAMEA4640836 | England, UK | 2018 | 12039 | 996 |
| WHO-X/H041 | PRJEB14020 | SAMEA2448468 | Japan | 2009 | 7363 | 226 |
| WHO-Y/F89 | PRJEB14020 | SAMEA2448469 | France | 2010 | 1901 | 16 |
| WHO-Z/A8806 | PRJEB14020 | SAMEA2796326 | Australia | 2013 | 7363 | 227 |
| YL201 | PRJNA560592 | SAMN20336117 | China | 2020 | 1600 | 2238 |
| ZJ19-F9 | PRJNA560592 | SAMN33303296 | China | 2019 | 1903 | 1143 |

| | | | | | | |
|---|---|---|---|---|---|---|
| ZJ20-230 | PRJNA560592 | SAMN33303299 | China | 2020 | 1600 | 2238 |
| ZJ20-9 | PRJNA560592 | SAMN33303302 | China | 2020 | 7827 | 5117 |

*This study

MLST, Multilocus Sequence Type; NG-STAR, *Neisseria gonorrhoeae* Sequence Typing for Antimicrobial Resistance

**Table S2.** Global ST8780 genomes used in Figure S1

| Sample | BioProject | BioSample | Country | Year | NG-STAR | *penA* allele |
|---|---|---|---|---|---|---|
| 16-191 | PRJEB47922 | SAMEA12276404 | Sweden | 2016 | 1207 | 2.002 |
| 16-599 | PRJEB47922 | SAMEA12278360 | Sweden | 2016 | 1309 | 2.008 |
| 554556 | PRJEB32435 | SAMEA5608448 | Norway | 2016 | 1664 | 2.008 |
| 4478STDY6638619 | PRJEB19989 | SAMEA5995066 | UK | 2015 | 1664 | 2.008 |
| 4478STDY6638666 | PRJEB19989 | SAMEA5995114 | UK | 2015 | 1664 | 2.008 |
| 4478STDY6638530 | PRJEB19989 | SAMEA5994980 | UK | 2015 | 1664 | 2.008 |
| 4478STDY6638533 | PRJEB19989 | SAMEA5994983 | UK | 2015 | 1664 | 2.008 |
| 4478STDY6638544 | PRJEB19989 | SAMEA5994994 | UK | 2015 | 1664 | 2.008 |
| 4478STDY6638550 | PRJEB19989 | SAMEA5994999 | UK | 2015 | 1664 | 2.008 |
| 4478STDY6638583 | PRJEB19989 | SAMEA5995033 | UK | 2015 | 1664 | 2.008 |
| TH22 | PRJEB47915 | SAMEA14294497 | Thailand | 2018 | 1664 | 2.008 |
| TH34 | PRJEB47915 | SAMEA14297518 | Thailand | 2018 | 1668 | 19.001 |
| 552703 | PRJEB32435 | SAMEA5608382 | Norway | 2016 | 2603 | 2.014 |
| 595716 | PRJEB32435 | SAMEA5608997 | Norway | 2016 | 3692 | 43.002 |
| AUSMDU00012468 | PRJNA520805 | SAMN10913575 | Australia | 2017 | 3692 | 43.002 |
| SAMEA6533518 | PRJEB32435 | SAMEA6533518 | Norway | 2016 | - | 9.001 |
| EEE019 | PRJNA776899 | SAMN22824079 | China | 2016 | - | 2.008 |
| JJJ022 | PRJNA776899 | SAMN22824261 | China | 2015 | - | 2.008 |
| EXNG1063 | PRJNA868503 | SAMN30248287 | Australia | 2017 | - | 2.008 |
| 16-191 | PRJEB47922 | SAMEA12276404 | Sweden | 2016 | 1207 | 2.002 |
| 16-599 | PRJEB47922 | SAMEA12278360 | Sweden | 2016 | 1309 | 2.008 |
| SGC-24-001* | PRJEB73456 | SAMEA116302675 | Scotland, UK | 2024 | 5998 | 60.001 |
| SGC-24-002* | PRJEB73456 | SAMEA116302676 | Scotland, UK | 2024 | 5998 | 60.001 |

*This study
NG-STAR, *Neisseria gonorrhoeae* Sequence Typing for Antimicrobial Resistance

**Table S3. Details of Scottish cases**

| Case | Year of Diagnosis | Sex | Age | Sexual Orientation | Anatomical Site | Symptomatic | HIV | *Chlamydia trachomatis* | Syphilis |
|---|---|---|---|---|---|---|---|---|---|
| SGC-23-001 | 2018 | M | 30-39 | Heterosexual | Genital | Yes | Neg | Neg | Neg |
| SGC-23-002 | 2022 | M | 20-29 | Heterosexual | Genital | Yes | Neg | Neg | Neg |
| SGC-23-003 | 2022 | F | 20-29 | Heterosexual | Genital | Yes | Neg | Neg | Neg |
| SGC-24-001 | 2024 | M | 20-29 | Heterosexual | Genital | Yes | Neg | Neg | Neg |
| SGC-24-002 | 2024 | F | 20-29 | Heterosexual | Genital | No | Neg | Neg | Neg |

F, female; M, male; Neg, negative

**Table S4.** Antimicrobial susceptibility testing results

| **Antimicrobial** | **SGC-23-001** | **SGC-23-002** | **SGC-23-003** | **SGC-24-001** | **SGC-24-002** |
|---|---|---|---|---|---|
| Azithromycin | 0.25 (WT) | >256.0 (nWT) | >256.0 (nWT) | 0.12 (WT) | 0.12 (WT) |
| Cefixime | 2.0 (R) | 2.0 (R) | 2.0 (R) | 3.0 (R) | 2.0 (R) |
| Ceftriaxone | 0.75 (R) | 0.19 (R) | 0.38 (R) | 1.0 (R) | 0.75 (R) |
| Ciprofloxacin | >1.0 (R) | >1.0 (R) | >1.0 (R) | >1.0 (R) | >1.0 (R) |
| Penicillin | 2.0 (R) | 0.5 (I) | 0.5 (I) | >2.0 (R) | >2.0 (R) |
| Spectinomycin | ≤64.0 (S) | ≤64.0 (S) | ≤64.0 (S) | ≤64.0 (S) | ≤64.0 (S) |
| Tetracycline | 4.0 (R) | >4.0 (R) | >4.0 (R) | >4.0 (R) | >4.0 (R) |

Antimicrobial minimum inhibitory concentrations are given in mg/L, breakpoints in parentheses as defined by EUCAST v13. nWT, non-wild type; R, resistant; S, susceptible; WT, wild type

**Table S5.** Genotypic resistance mechanisms

| **Marker** | **Drug affected** | **SGC-23-001** | **SGC-23-002** | **SGC-23-003** | **SGC-24-001** | **SGC-24-002** |
|---|---|---|---|---|---|---|
| 23S rDNA A2059G | AZM | - | Detected | Detected | - | - |
| Mosaic *penA* | CFM/CRO/PEN | Detected | Detected | Detected | Detected | Detected |
| *gyrA* S91F & D95A | CIP | Detected | Detected | Detected | Detected | Detected |
| *parC* S87N | CIP | - | - | - | Detected | Detected |
| *parC* S87R | CIP | Detected | Detected | Detected | - | - |
| *mtrR* promoter deletion | AZM/PEN/TET[1] | Detected | - | - | Detected | Detected |
| $bla_{TEM}$ | PEN | - | - | - | Detected | Detected |
| *mtrR* A39T | PEN | - | Detected | Detected | Detected | Detected |
| *ponA1* L421P | PEN | Detected | Detected | Detected | Detected | Detected |
| *porB1b* G120K & A121D | PEN | Detected | - | - | Detected | Detected |
| *rpsJ* V57M | TET | Detected | Detected | Detected | Detected | Detected |
| *tetM* | TET | - | Detected | Detected | Detected | Detected |

[1]*mtrR* promoter deletion alone is not sufficient for azithromycin resistance (6)
AZM, azithromycin; CFM, cefixime; CIP, ciprofloxacin; CRO, ceftriaxone; PEN, penicillin; TET, tetracycline. No spectinomycin resistance determinants were detected.

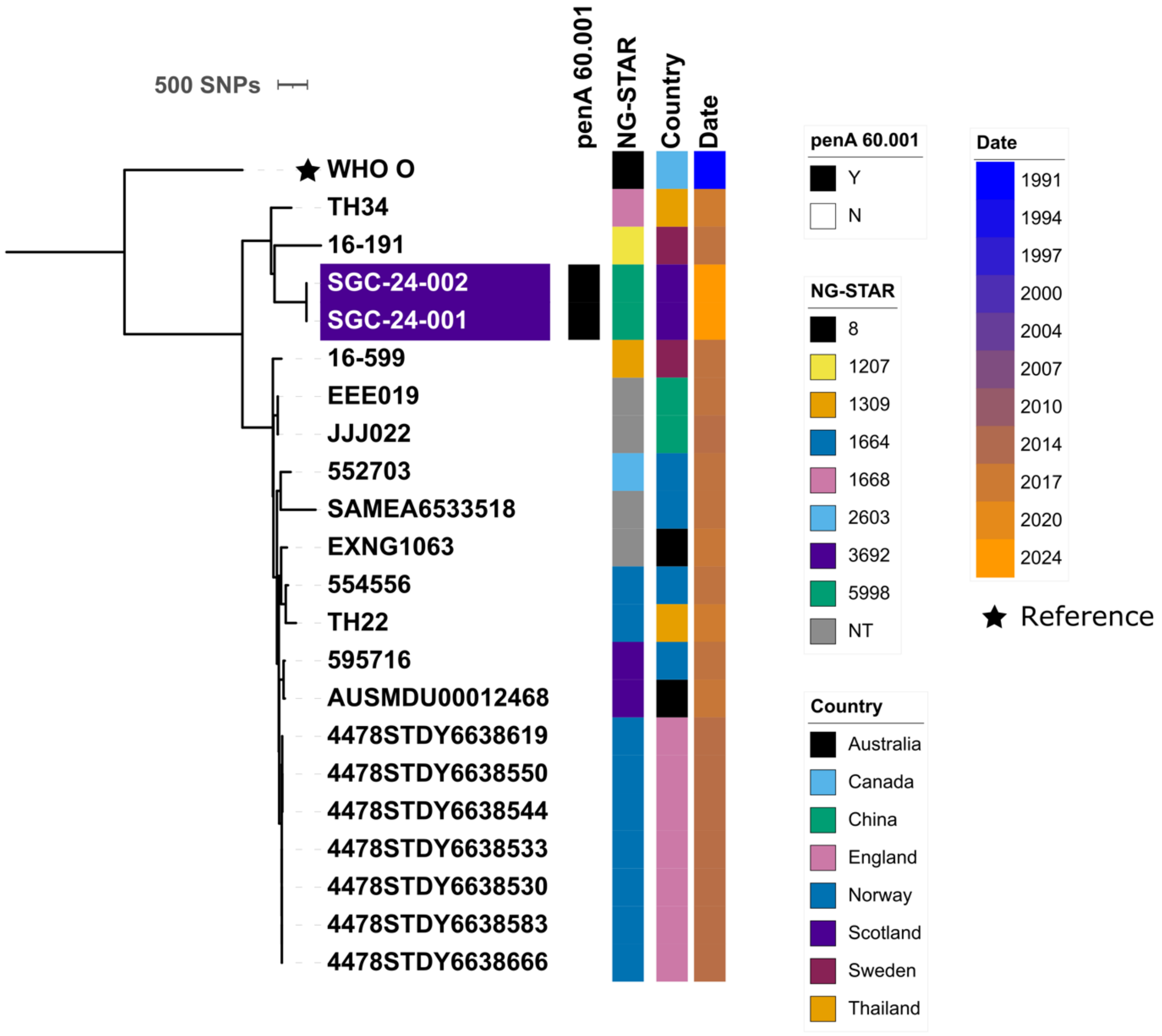


**Figure S1** Maximum-likelihood phylogeny of 21 ST8780 genomes. Scottish genomes are highlighted in purple. Metadata bars coloured as per figure legend.
